# Prospective Validation to Predict Radiofrequency Lesion Size During High-Power and Short-Duration Ablation Using a Novel Index Formula Incorporating Contact Force, Radiofrequency Power and Application Time in a Swine Beating Heart Model

**DOI:** 10.1101/2025.03.19.25324285

**Authors:** Masafumi Sugawara, Atsushi Ikeda, Assaf Govari, Zach Bubar, Tushar Sharma, Christopher T. Beeckler, Arwa Younis, Chadi Tabaja, Ayman A. Hussein, Shady Nakhla, Pasquale Santangeli, Walid I. Saliba, Oussama M. Wazni, Warren M. Jackman, Hiroshi Nakagawa

## Abstract

**Background:** During radiofrequency (RF) ablation, lesion size increases with increasing contact force (CF), RF power and application time. The effects of CF and RF power on lesion size during high-power and short-duration (HP-SD) ablation have not been well-determined. This study aimed to, during HP-SD ablation: 1) examine the relationship between lesion size and CF, RF power and time, and 2) prospectively validate the ability of a novel logarithmic formula, incorporating CF, RF power and time (Force-Power-Time-Index, FPTI, gram x Watt x sec) to predict lesion size using a swine beating heart model.

**Methods:** Eight closed-chest swine were studied. A 7.5Fr CF ablation catheter with a 3.5mm irrigated-tip electrode containing 6 surface thermocouples (Qdot-Micro) was positioned in the right and left ventricles. In five swine (Phase1-Study), RF was delivered at ≤90Watts (modulated to maintain the surface electrode temperature<65°C) for 4sec to 103 ventricular sites with various CF (range 5-54*g*). Swine were sacrificed and lesion size was measured. A new logarithmic FPTI-Formula was created based on the relationship between lesion depth and CF, power and time. In the prospective validation study using the remaining three swine (Phase2-Study), RF(90W) was delivered for 4 sec at 72 sites with FPTI-Formula predicted lesion depths of 2-6mm. Actual lesion depth was compared to the predicted lesion depth.

**Results:** In the Phase1-study, there was a close relationship between lesion depth and the product of Force x Power x Time (R=0.711, *p*<0.0001), creating a novel logarithmic FPTI-Formula to predict lesion depth. In the Phase2-study, lesion depth predicted by the FPTI-Formula correlated highly with actual lesion depth (1.9-6.1mm), with ±1mm accuracy in 68/72(94%) lesions (R=0.867, *p*<0.0001). No steam pop or thrombus formation occurred.

**Conclusion:** During HP-SD ablation, the new FPTI-Formula prospectively predicted lesion depth with high accuracy while the surface electrode temperature control prevented steam pop and thrombus formation.

## INTRODUCTION

In ablation for patients with atrial fibrillation (AF) and macro-reentrant atrial tachycardia, it is desired to create continuous and transmural ablation lesions while minimizing the risk of complications, including collateral tissue injury, steam pops, and thrombus formation. Preclinical studies have shown that high-power and short-duration (HP-SD) radiofrequency (RF) ablation may create wider and shallower lesions, facilitating effective and safe AF ablation with a shorter procedure time.^1–3^ However, recent clinical studies on pulmonary vein (PV) isolation using HP-SD RF ablation have shown significant variability in the efficacy of PV isolation (first-pass PV isolation rates ranging from 18% to 83% and acute reconnection rates ranging from 4% to 32%), as well as the occurrence of esophageal injury.^4–10^

During conventional RF ablation (25-45 Watts) using a saline-irrigated electrode, ablation lesion size (depth and diameter) increases with increasing three dominant variables: contact force (CF), RF power, and application time.^11–15^ Based on the pre-clinical canine study comparing lesion depth of various combinations of CF (5-50*g*), RF power (25-45 Watts), and application time (15-60 sec), a logarithmic formula (Force-Power-Time Index, Ablation Index) has been developed to predict lesion depth during RF ablation.^16,17^ The Ablation Index indicates when to terminate the RF application to obtain lesions in the range of 3-9 mm depth with high prediction accuracy in beating canine hearts. Several clinical studies using the Ablation Index have demonstrated its usefulness in controlling lesion size, including a high incidence of PV isolation with the first PV encirclement and high durability of PV isolation.^17–20^

Nevertheless, the effects of CF and RF power on lesion size during high-power (50-90 Watts) and short-duration (4-5 sec) applications have not been well determined. The purpose of the present study is, during very high power (≤90 Watts) and very short duration (4 sec) ablation in a swine heart model: 1) to examine the relationship between lesion size (depth and diameter) and CF, RF power and application time; 2) to develop a novel logarithmic formula based on these data to predict lesion size, i.e., Force-Power-Time-Index (FPTI) – ***“Phase 1 study”*;** and 3) to prospectively validate the accuracy of the new Force-Power-Time Index Formula to predict lesion depth during HP-SD ablation in a swine beating heart study – ***“Phase 2 study.”***

## METHODS

The data supporting this study’s findings are available from the corresponding author upon reasonable request.

### High-Power and Short-Duration (90 Watts/4 sec) Ablation System

A 7.5Fr ablation catheter had a 3.5 mm irrigated-tip electrode containing 66 tiny irrigation holes around the electrode, a CF sensor, three micro-recording electrodes on the distal end, and six thermocouples located only 75μm from the surface, three at the distal end and three at the proximal end of the electrode (Qdot-Micro, Johnson &Johnson MedTech, Inc., Irvine CA, **Fig 1**). The symmetrical distribution of the proximal and distal thermocouples improved the accuracy of recording the surface electrode temperature in both perpendicular and parallel electrode-tissue orientations.^21^ In order to minimize the risk of thrombus formation, the irrigation holes at the distal and proximal ends of the ablation electrode were angled distally and proximally, respectively, improving cooling at the distal and proximal electrode edges during high-power RF applications (**Fig 1**).^21^ A multi-channel RF generator (nGEN, Johnson &Johnson MedTech) delivered RF power to 90 Watts.

**Figure 1.**
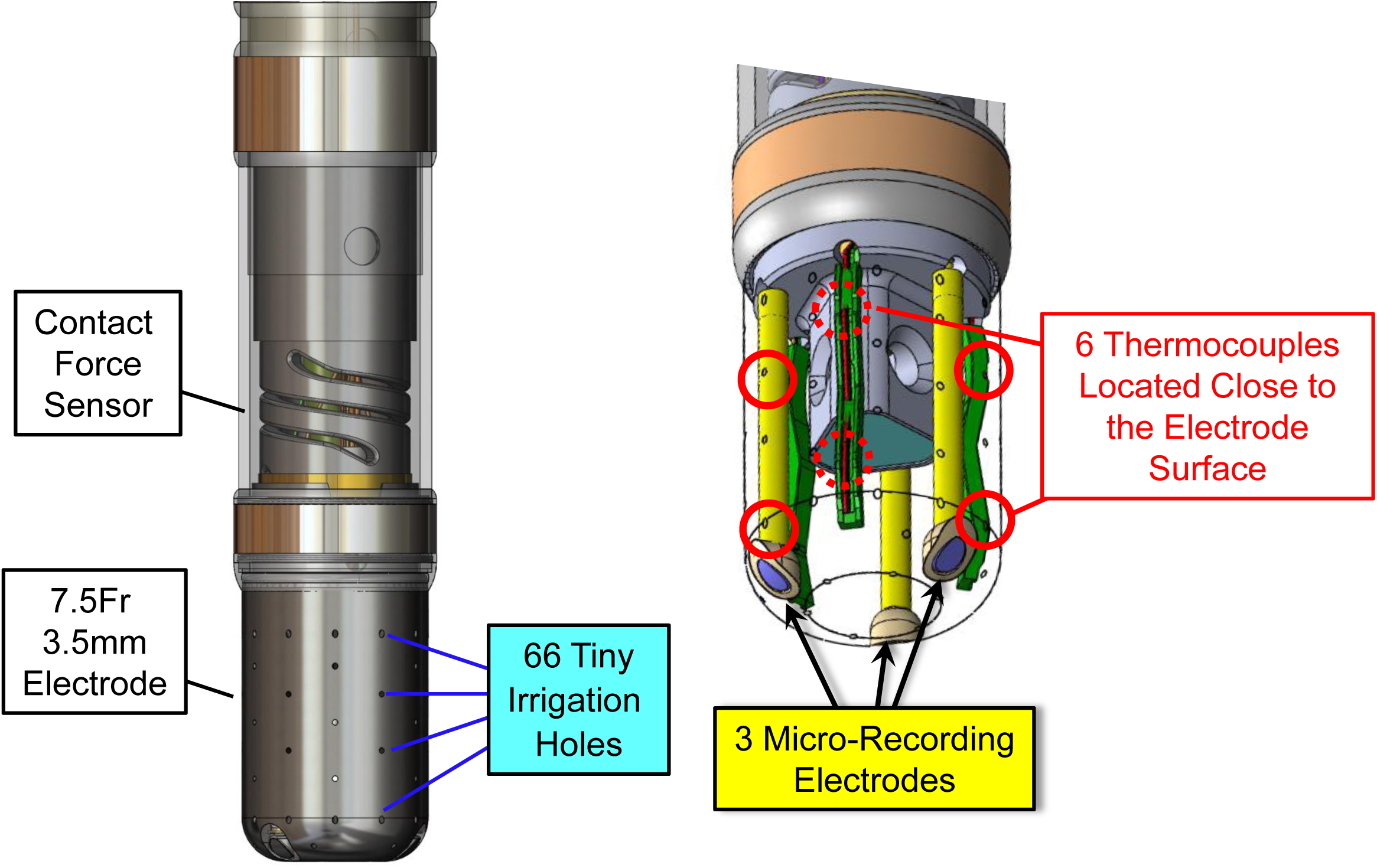
Schematic representation of the study catheter for high-power (90 Watts) and short-duration (4 sec), HP-SD, RF ablation. **See text for details.**

### Experimental Preparation

The experimental protocol was approved by the Committee on the Use and Care of Animals at the Cleveland Clinic Foundation, Cleveland, OH. Eight Yorkshire swine weighing 68 to 80 kg were anesthetized with isoflurane and ventilated mechanically. Heparin (10,000-12,000 IU) was administered intravenously with additional doses, as necessary to maintain the activated clotting time at ≥300 sec. A 6Fr decapolar catheter was inserted into the right internal jugular vein and advanced into the coronary sinus (CS). An 8.5Fr ultrasound catheter (AcuNav, Acuson) was inserted into the left femoral vein and advanced into the right atrium for intracardiac echocardiography (ICE). Transeptal puncture was performed under ICE and fluoroscopic guidance. A deflectable transeptal sheath (Vizigo, Johnson &Johnson MedTech) was inserted into the right femoral vein and advanced into the left atrium (LA) and left ventricle (LV). An anatomical shell of the LV chamber was created using a multi-electrode mapping catheter (Octaray, Johnson &Johnson MedTech) and the CARTO3 mapping system (Johnson &Johnson MedTech). The Octaray mapping catheter was exchanged for the ablation catheter (Qdot-Micro). The ablation catheter was initially positioned centrally within the LA without electrode-tissue contact (confirmed by ICE) to calibrate the CF-sensor to 0*g* (baseline non-contact value). The ablation catheter was then advanced into the LV for ablation. Before ablation, the decapolar catheter in the CS was re-positioned into the right ventricular (RV) apex for pacing. After LV ablation, the Octaray catheter was moved to the RV to generate the RV anatomical shell, and RV ablation was performed.

### Phase 1 Study: Examination of the Relationship Between Contact Force, RF Power and Lesion Size During HP-SD Ablation

#### 1) Ablation Protocol

During RV apical pacing at a cycle length of 400ms, high-power and short-duration (HP-SD) ablation was performed by delivering RF power at up to 90 Watts, modulated down to maintain a surface electrode temperature of <65°C, for 4 sec between the ablation electrode and a skin patch (positioned on the posterior chest) during saline irrigation of 8 ml/min (**Fig 2A**). The CF, RF-power, impedance, and surface electrode temperature were continuously monitored and recorded. The ablation sites were stored on the CARTO3 mapping system and were sufficiently separated to be accurately identified during lesion assessment (**Fig 2B**).

**Figure 2.**
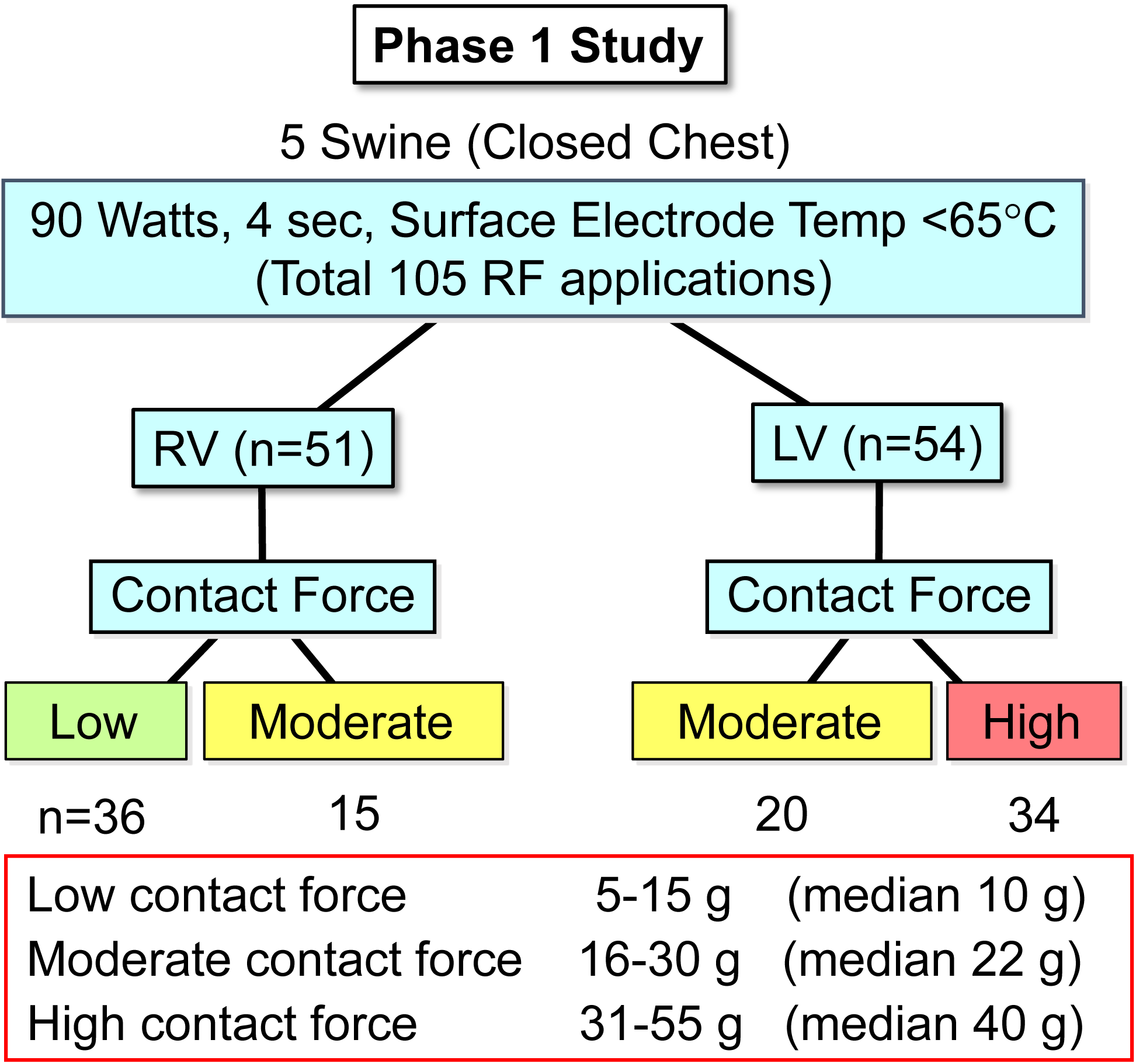
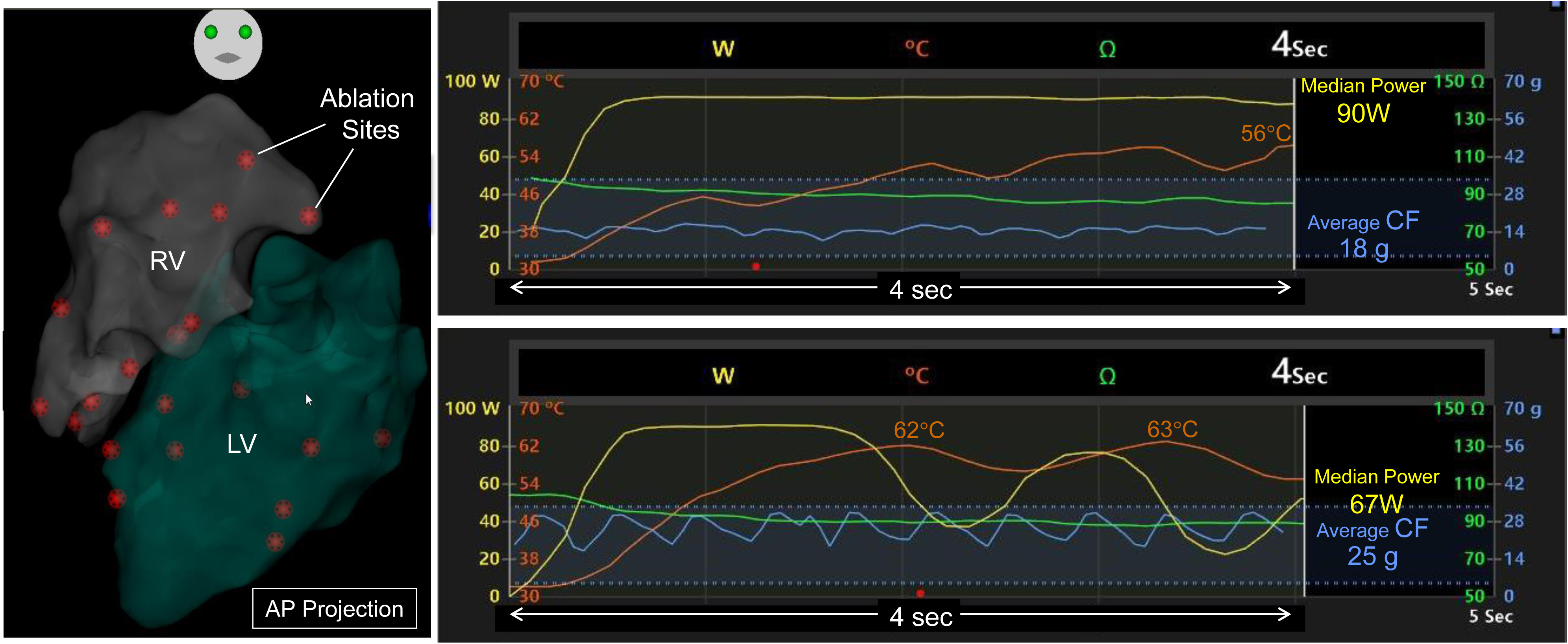
Protocol of the Phase 1 study on HP-SD RF ablation and RF parameters. **A**. In the first five swine, HP-SD RF ablation was performed at 51 RV and 54 LV sites using at three different CF levels: 1) low CF (range 5-15*g*); 2) moderate CF (range 16-30*g*); and 3) high CF (range 31-55*g*). **B.** RF parameters of CF (light blue), RF power (yellow), RF time (white), surface electrode temperature (orange, displaying the highest temperature of the six thermocouples) and impedance (green) are recorded during HP-SD ablation with either full RF power application due to peak electrode temperature of 56°C, i.e., <65°C (median power of 90 Watts, right upper panel) or RF power modulation (median power of 67 Watts) to maintain the surface electrode temperature of <65°C (right lower panel). CARTO image in the anterior-posterior (AP) projection (left panel) showing each ablation site (brown tag) in the right ventricle (RV) and left ventricle (LV).

In the first 5 of the eight swine, RF ablation was performed at 10-12 separate sites in the LV and 10-11 separate sites in the RV in each animal at three different CF levels: 1) low CF (range 5-15*g*); 2) moderate CF (range 16-30*g*); and 3) high CF (range 31-55*g*, **Fig 2A**).

#### 2) Measurements of Lesion Size

After the ablation procedure, 250 ml of 1% triphenyl tetrazolium chloride (TTC) was intravenously administered. The animals were sacrificed, and the hearts were excised and fixed in 10% formalin. The hearts were then sectioned at 2-3 mm slices perpendicular to the intraventricular groove from the apex to the base (**Fig 3A**). For each lesion, the maximum depth (A), maximum diameter (B), depth at the maximum diameter (C), and surface diameter (D) were measured by two investigators blinded to the ablation parameters (**Fig 3B**). Lesion volume was calculated as volume = (1/6)×π×(A×B^2^+C×D^2/2^). ^21, 22^

**Figure 3.**
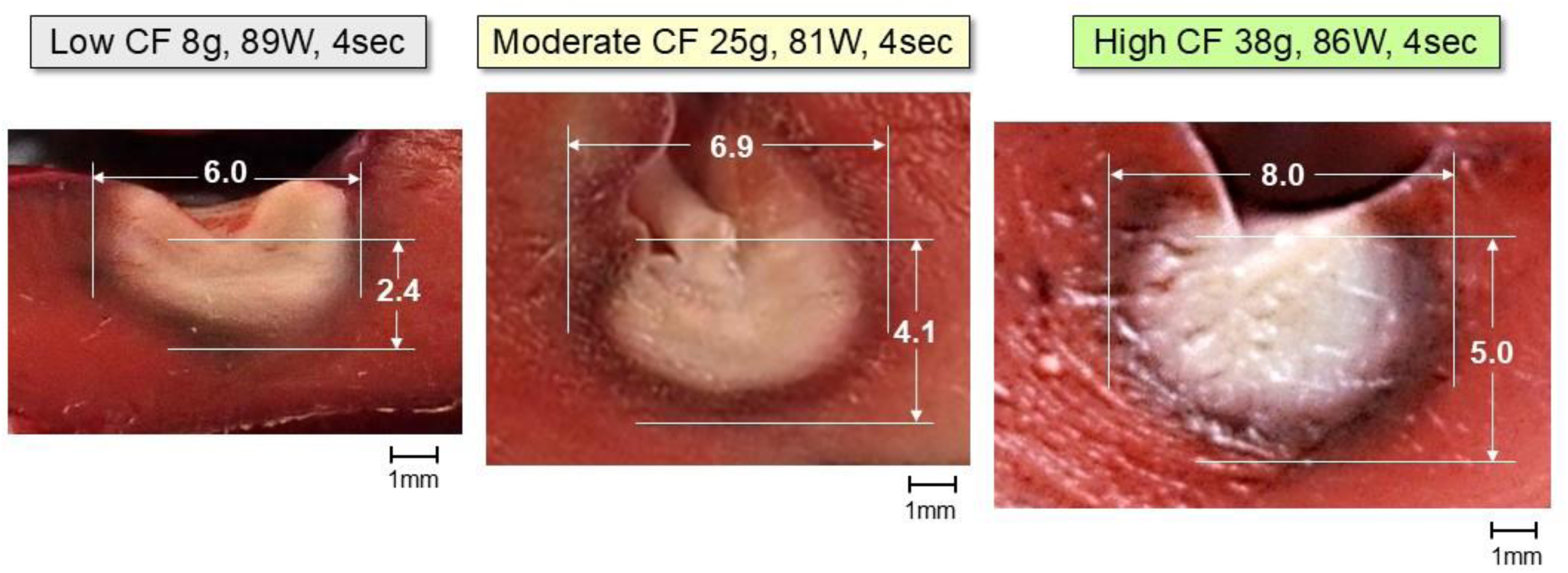
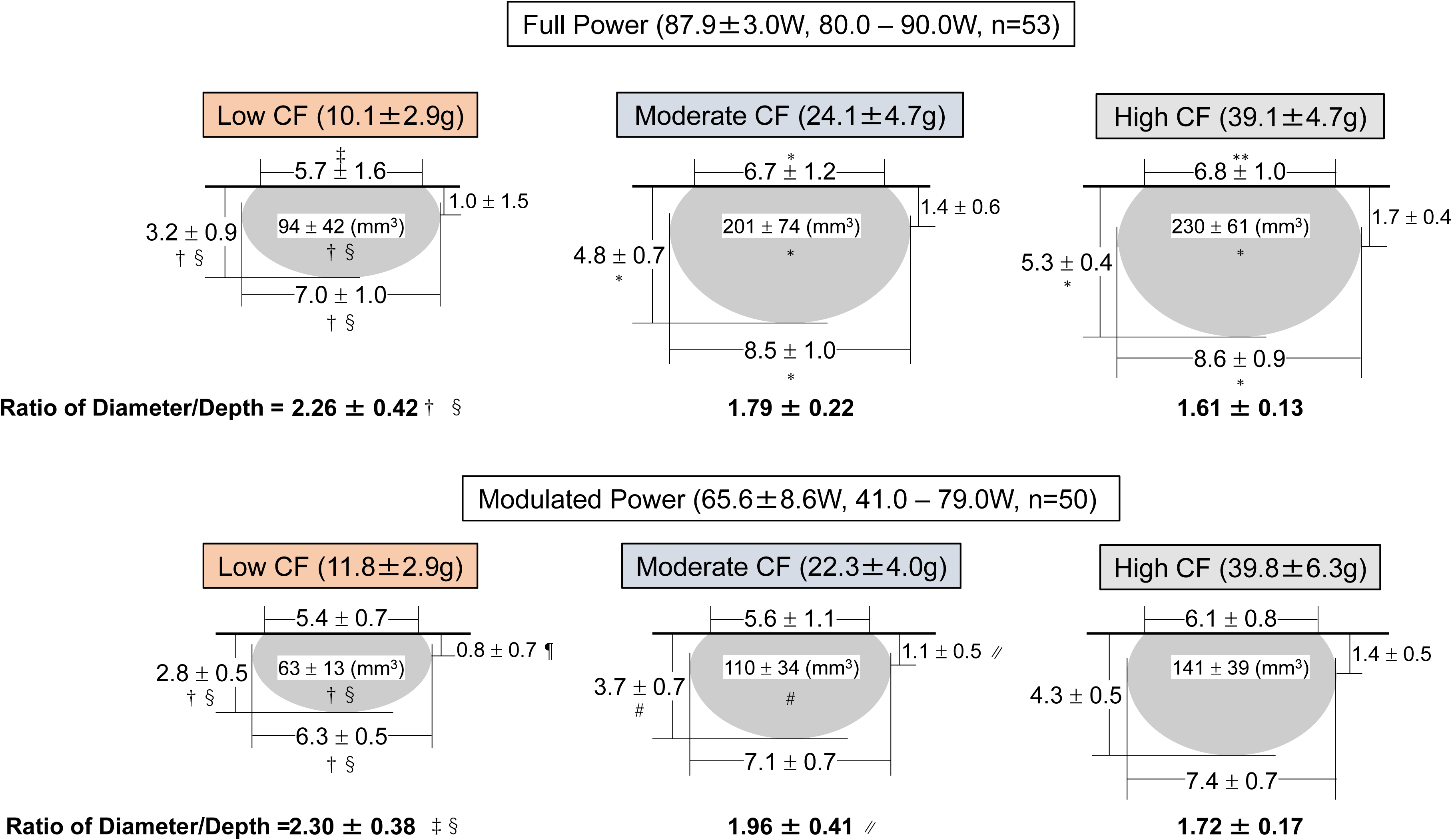
Ablation lesion size produced by HP-SD ablation with low, moderate, and high contact force (CF). **A.** Photographs of lesions produced by HP-SD ablation with full RF power (defined as median RF power of ≥80 Watts) applications. Increasing average CF of 8*g*, 25*g*, and 38*g* results in greater lesion depths of 2.4 mm, 4.1 mm, and 5.0 mm, respectively, and greater diameters of 6.0 mm, 6.9 mm, and 8.0 mm, respectively. **B.** Diagram of lesion dimensions for HP-SD ablation with full RF power applications (87.9 ± 3.0 Watts, n=53, upper panel) and modulated RF power applications (65.6 ± 8.6 Watts, n=50, lower panel) at low, moderate and high average CF, respectively. Lesion measurements (mm, mean ± SD) include maximum depth (number left of each figure), maximum diameter (bottom of each figure), surface diameter (top of each figure), and depth at maximum diameter (right of each figure), calculated lesion volume (mm^3^, inside each figure) and ratio of diameter/depth. See text for details. †*p*<0.01 for low CF vs moderate CF; ‡*p*<0.05 for low CF vs moderate CF; §*p*<0.01 for low CF vs high CF; ¶*p*<0.05 for low CF vs high CF, #*p*<0.01 for moderate CF vs high CF; ∥*p*<0.05 for moderate CF vs high CF; \**p*<0.01 for full power vs modulated power;\*\**p*<0.05 for full power vs modulated power.

#### 3) Data Analysis for RF Ablation Parameters and Lesion Size

The relationships between lesion size (maximum depth and diameter) and median RF power, average CF, impedance decrease, and surface electrode temperature were examined (**Figs 4A-C**).

**Figure 4.**
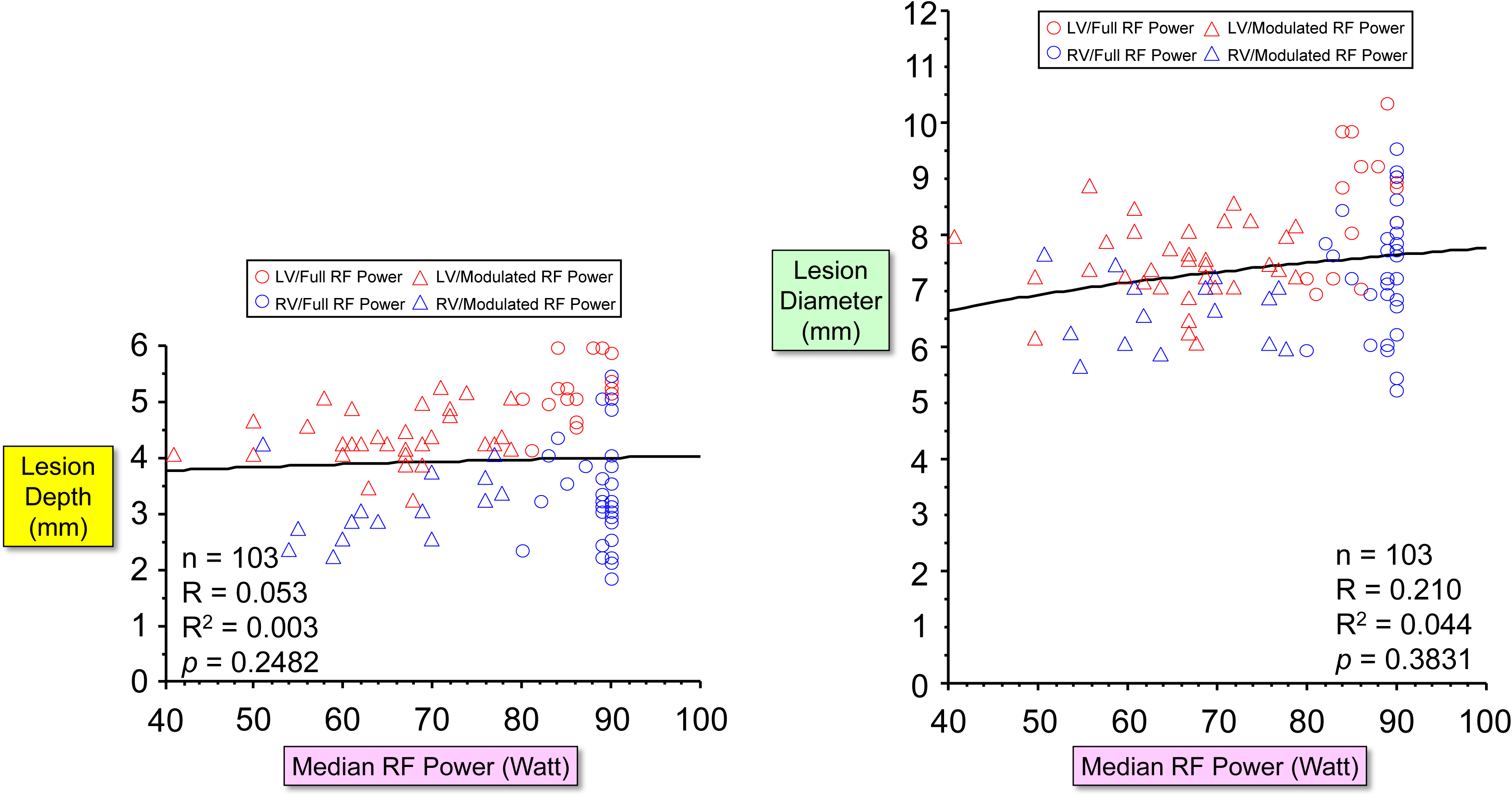
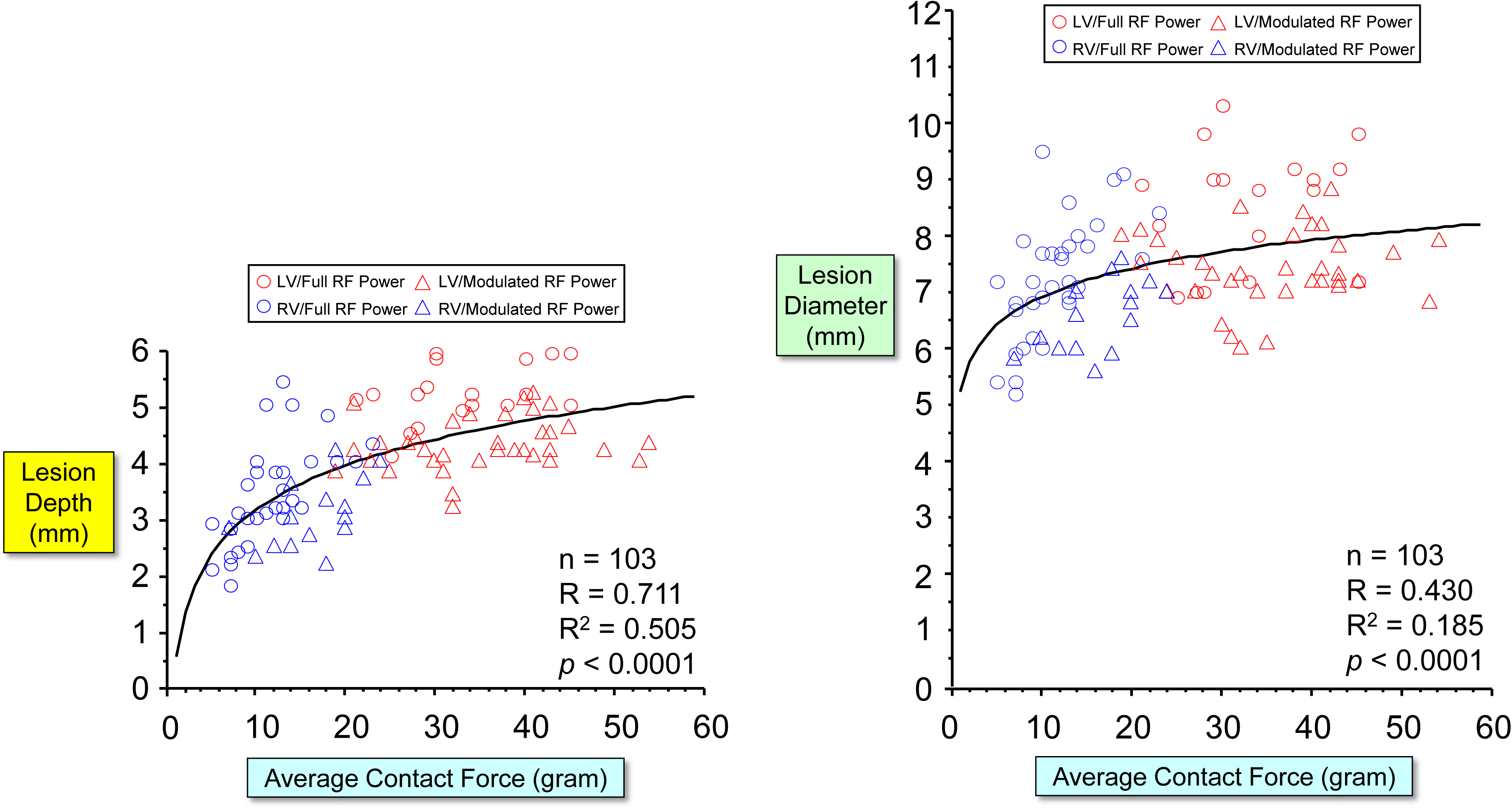
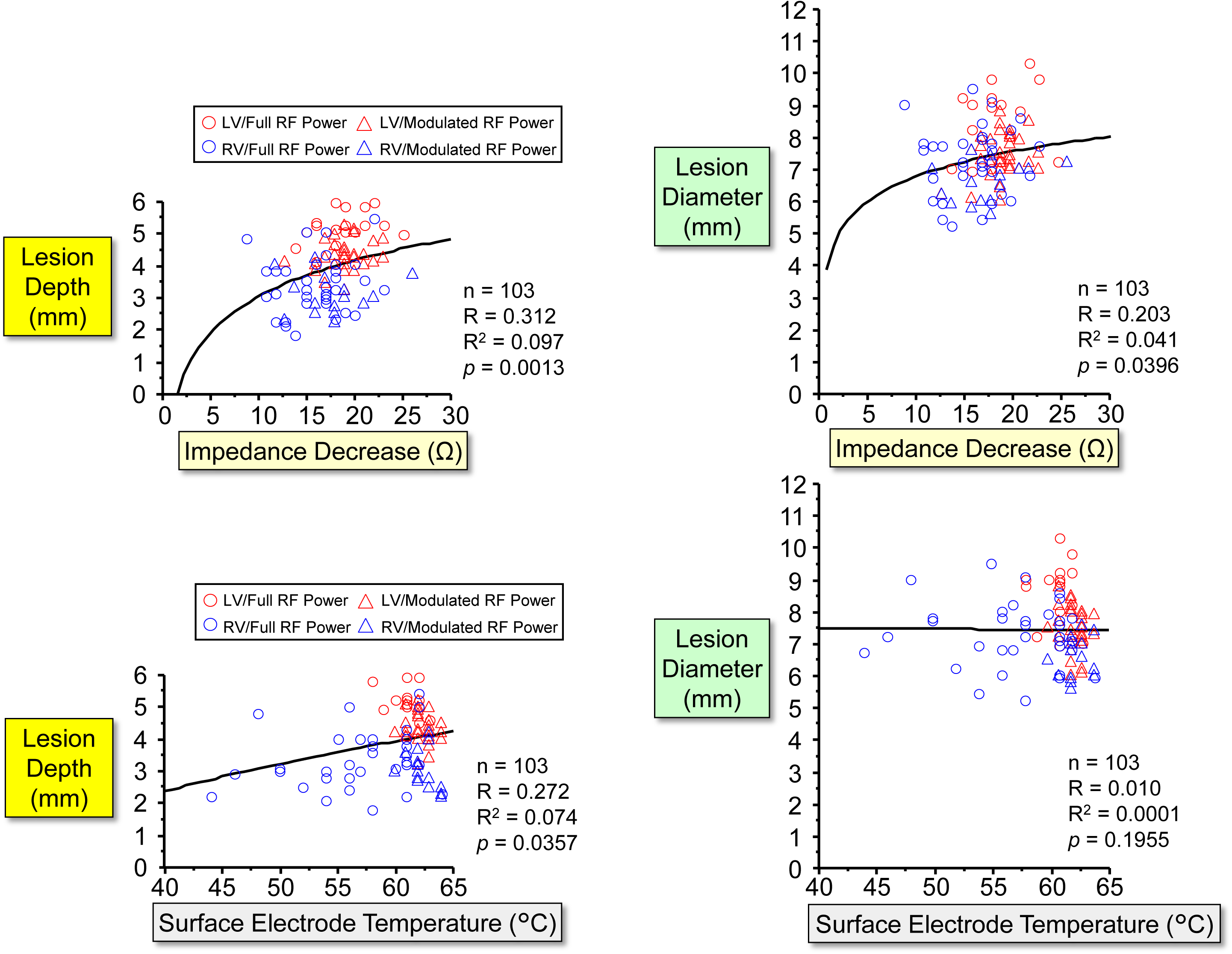
Relationships between median RF power, average contact force, impedance decrease, surface electrode temperature, and lesion size. **A.** No significant correlation between median RF power and lesion depth (left panel) and lesion diameter (right panel). **B.** Significant correlations between average CF and lesion depth (R=0.711, *p*<0.0001, left panel) and lesion diameter (R=0.430, *p*<0.0001, right panel). **C.** There are poor correlations between the amount of impedance decrease and lesion depth and diameter (R=0.312, *p*=0.0013 and R=0.203, *p*=0.0396, respectively, upper panels). There is a poor or no correlation between the maximum surface electrode temperature and lesion depth or lesion diameter (R=0.272, *p*=0.0357 and R=0.010, *p*=0.1955, respectively, lower panels). Red circle: LV ablation with full RF power applications, red triangle: LV ablation with modulated RF power applications, blue circle: RV ablation with full RF power applications, and blue triangle: RV ablation with modulated RF power applications,

The extent of myocardial trabeculation at the ablation sites was classified into three grades (**Fig 5A**). The relationships between the trabeculation grade and the degree of RF power modulation (which was required to maintain the surface electrode temperature of <65°C) and between the trabeculation grade and the initial impedance measured at the onset of RF application were analyzed (**Fig 5B**).

**Figure 5.**
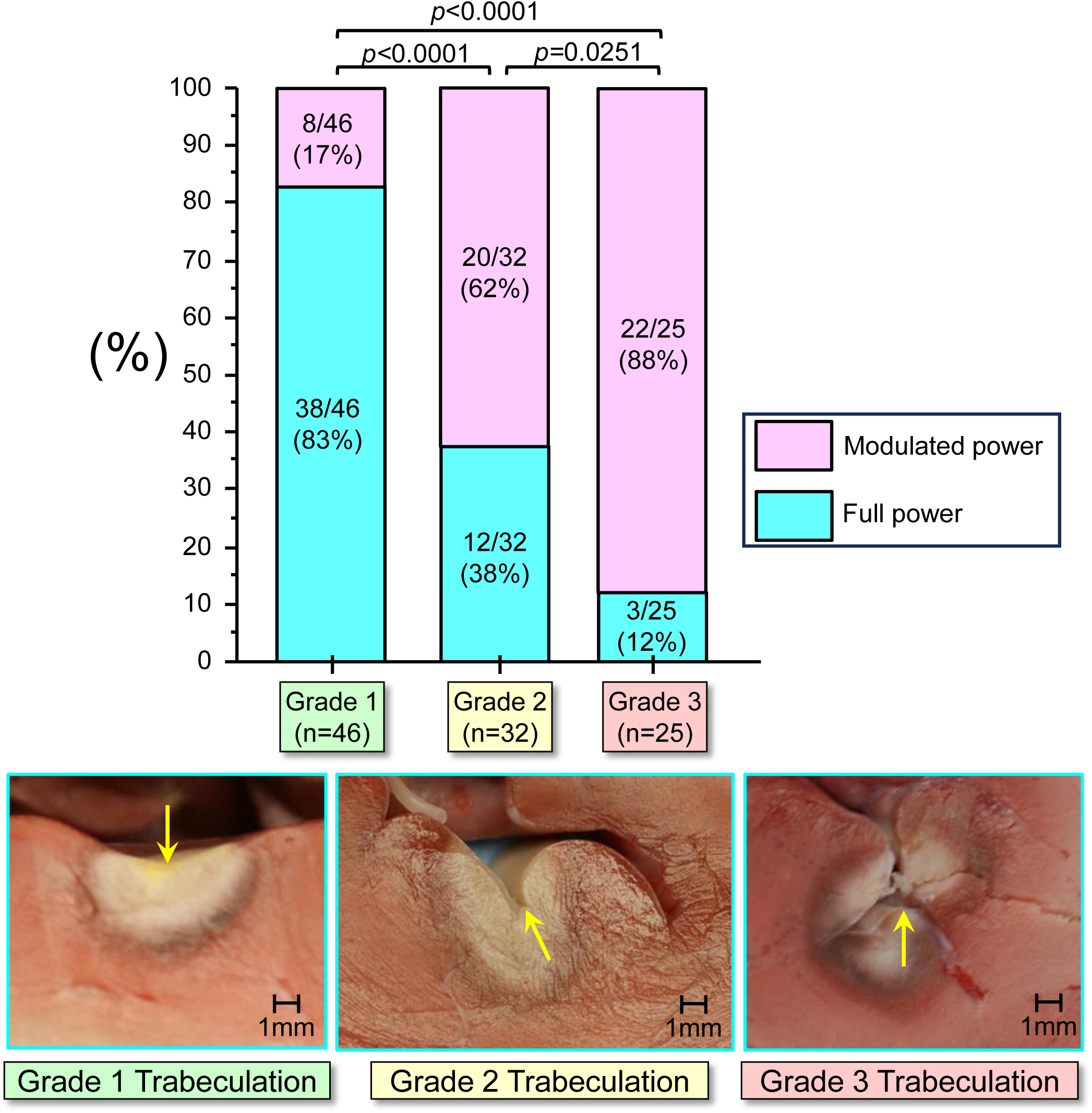
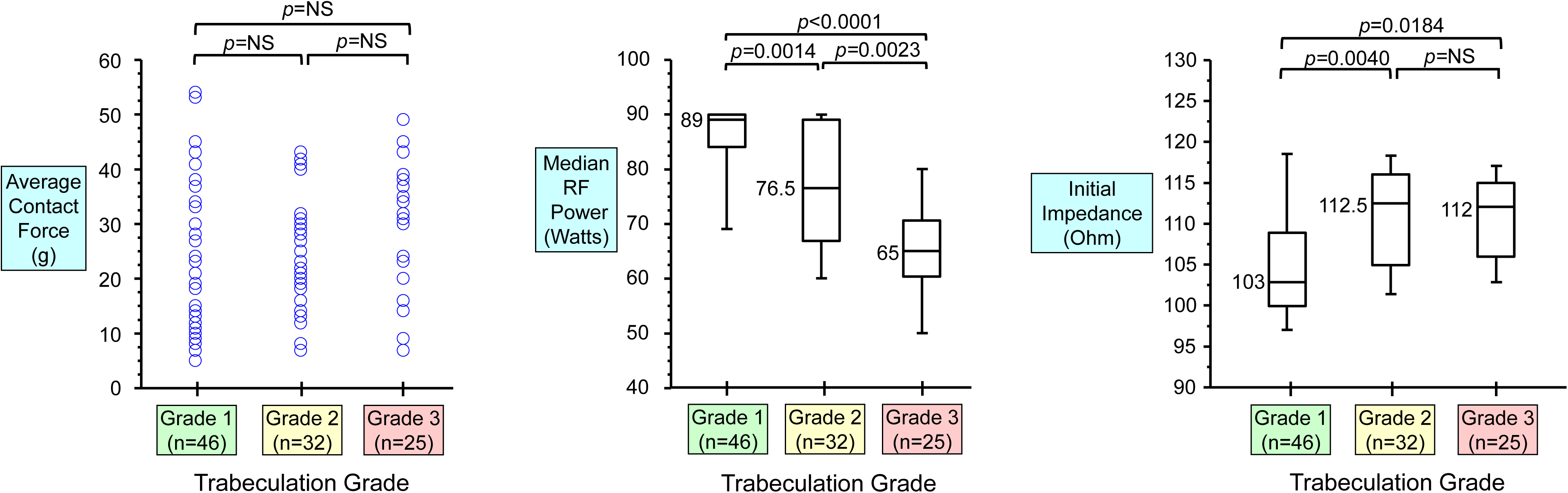
Relationship RF power modulation and the myocardial trabeculation grade. **A.** The extent of myocardial trabeculation at the ablation site is classified into three grades, grade 1 (no trabeculation), grade 2 (moderated trabeculation) and grade 3 (high trabeculation, an ablation electrode seems to be embedded deep within the trabecular grooves). The yellow arrow indicates the estimated position of the ablation tip electrode. As the trabeculation grade increases, the significantly higher incidence of RF power modulation to maintain the surface electrode temperature of < 65°C is observed: 8/46 (17%) at grade 1 ablation sites, 20/32 (62%) at grade 2 ablation sites, and 22/25 (88%) at grade 3 ablation sites. **B.** There is no significant difference in average CF among the three grades of trabeculation at ablation sites (left panel). However, the median RF power to maintain the surface electrode temperature of <65°C is significantly lower as the trabeculation grade increases (grade 1 trabeculation: 89 Watts, grade 2 trabeculation: 76.5 Watts, and grade 3 trabeculation: 65 Watts, center panel). The initial impedance is significantly lower at grade 1 trabeculation sites (median 103 Ohms) than grade 2 and 3 trabeculation sites (112.5 Ohms and 112 Ohms, respectively, right panel). The box plots show the median (50th percentile), 10th, 25th, 75th, and 90th percentile values.

In order to estimate the amount of RF energy delivered to the myocardium, a product of average CF (gram) x median RF power (Watt) x application time (sec), (Force-Power-Time Index: FPTI) was calculated with each RF application. The relationship between lesion size (depth and diameter) and FPTI value was examined (**Fig 6A**). A logarithmic formula (***FPTI Formula***) based on the relationship between lesion size and FPTI was developed, and the relationship between the formula’s estimated lesion size and actual size was analyzed (**Fig 6B**).

**Figure 6.**
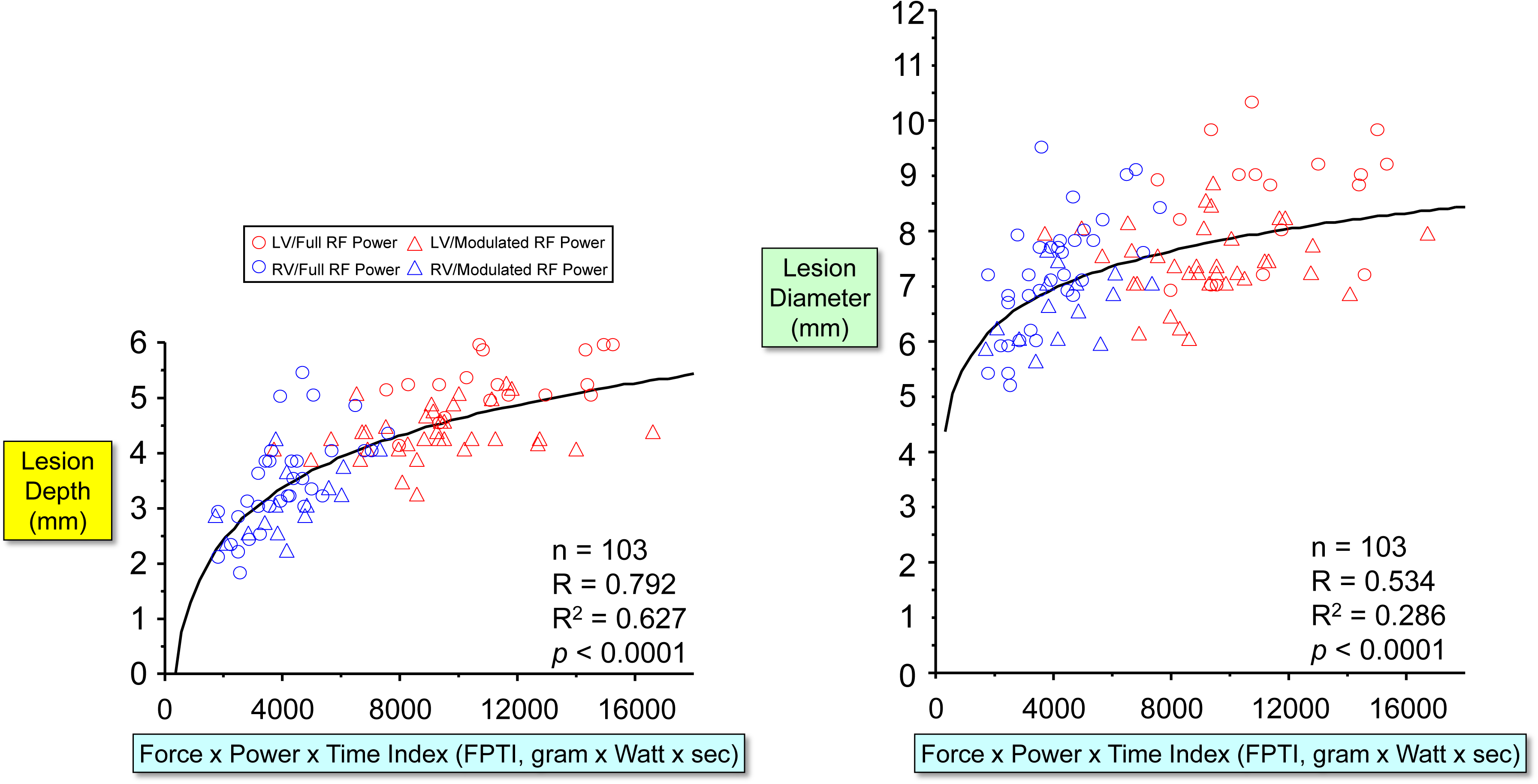
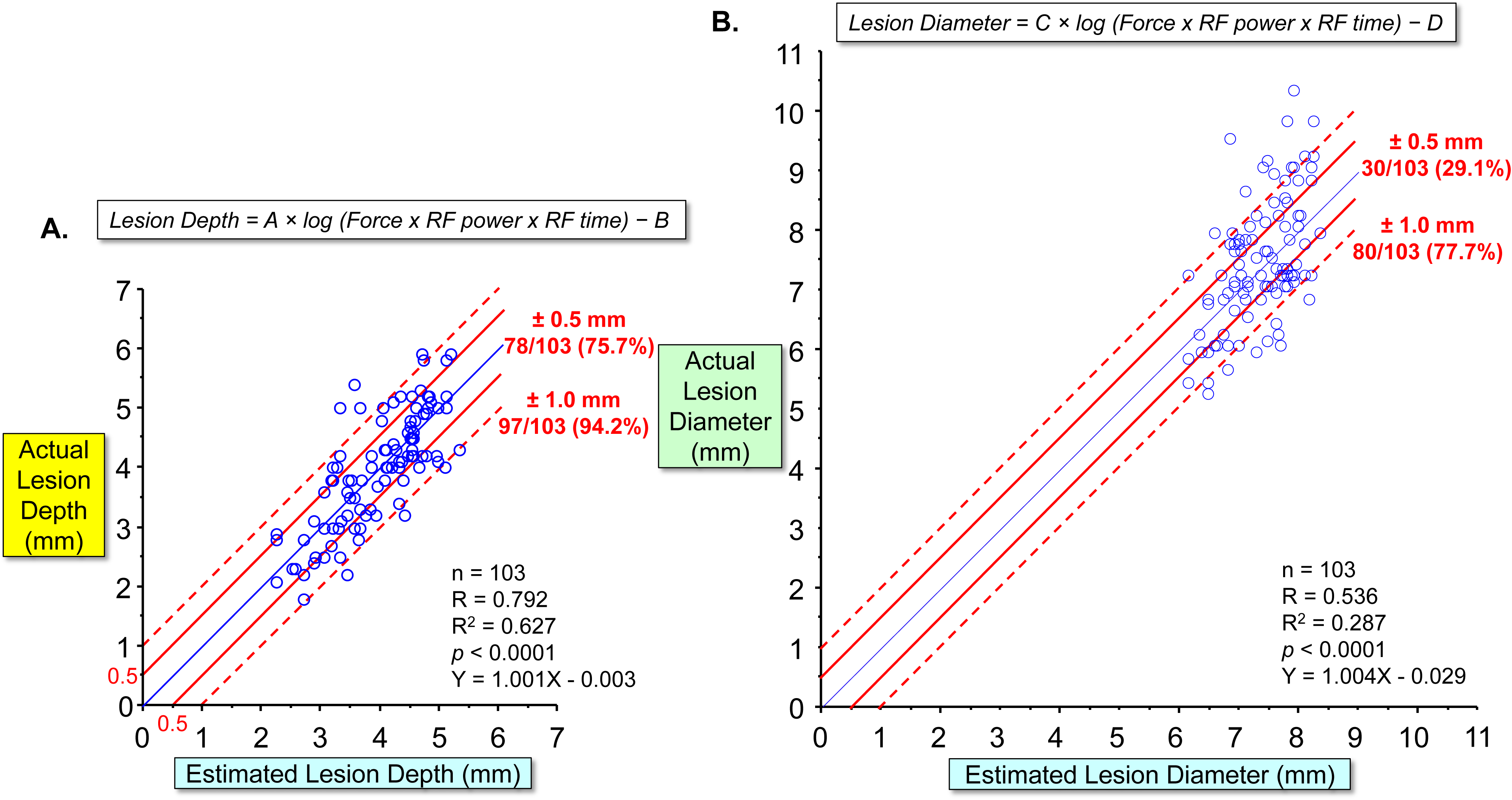
Relationship between a product of average CF (gram) x median RF power (Watt) x application time (sec), Force-Power-Time Index (FPTI) and lesion size. **A.** Significant correlations between Force-Power-Time Index (FPTI) and lesion depth and diameter (R=0.792, *p*<0.0001 and R=0.534, *p*<0.0001, respectively). **B.** The logarithmic formulas are created based on the relationships between the FPTI values and lesion depth and diameter, respectively: 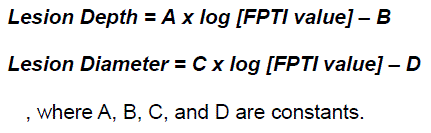 The difference between the estimated lesion depth by the formula and actual lesion depth was within ±0.5 mm in 78/103 (75.7%) lesions and within ±1.0 mm in 97/103 (94.2%) lesions (**left panel**) and the difference between the estimated lesion diameter and actual diameter was within ±0.5 mm in 30/103 (29.1%) lesions and within ±1.0 mm in 80/103 (77.7%) lesions (**right panel**).

### Phase 2 Study: Prospective Validation of FPTI Formula to Predict Lesion Depth During HP-SD Ablation

#### 1) Ablation Protocol

In the remaining three swine, HP-SD ablation was performed by delivering 90 Watts for 4 sec with RF power modulation to maintain the surface electrode temperature of <65°C at 72 separate ventricular sites, with the FPTI-Formula’s predicted lesion depths of 2.0-3.0mm, 3.1-4.0mm or 4.1-5.0 mm with the average CF ranging 4-26*g* in the RV (n=36), and the predicted lesion depths of 3.1-4.0 mm, 4.1-5.0 mm or 5.1-6.0 mm with the average CF ranging 26-56*g* in the LV (n=36), respectively (**Fig 7**).

**Figure 7.**
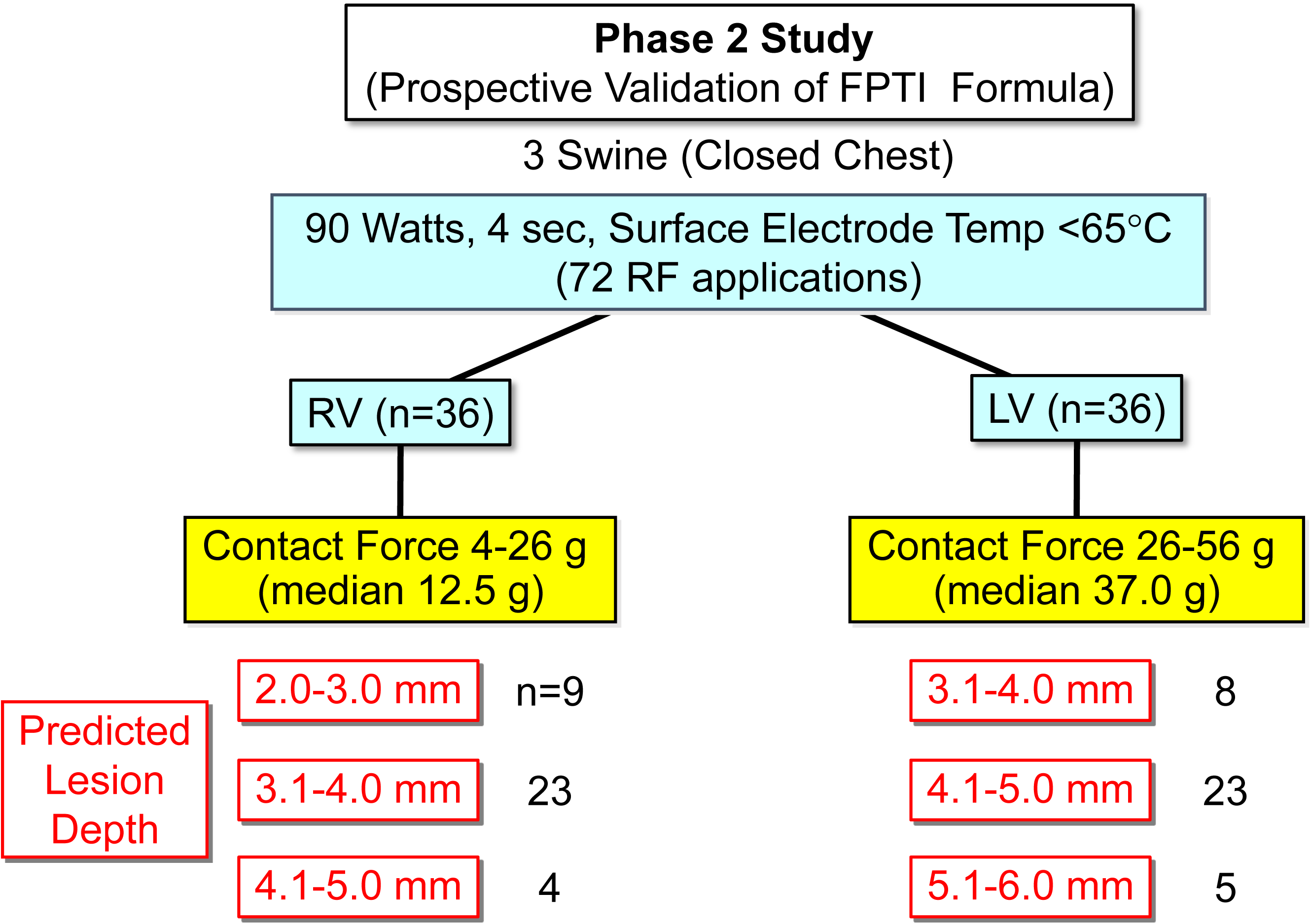
Protocol of the Phase 2 prospective validation study to test the ability to predict lesion depth using the FPTI formula during HP-SD ablation. See text for details.

#### 2) Measurements of Lesion Size

After ablation, lesion size was measured as described above (**Fig 8A**).

**Figure 8.**
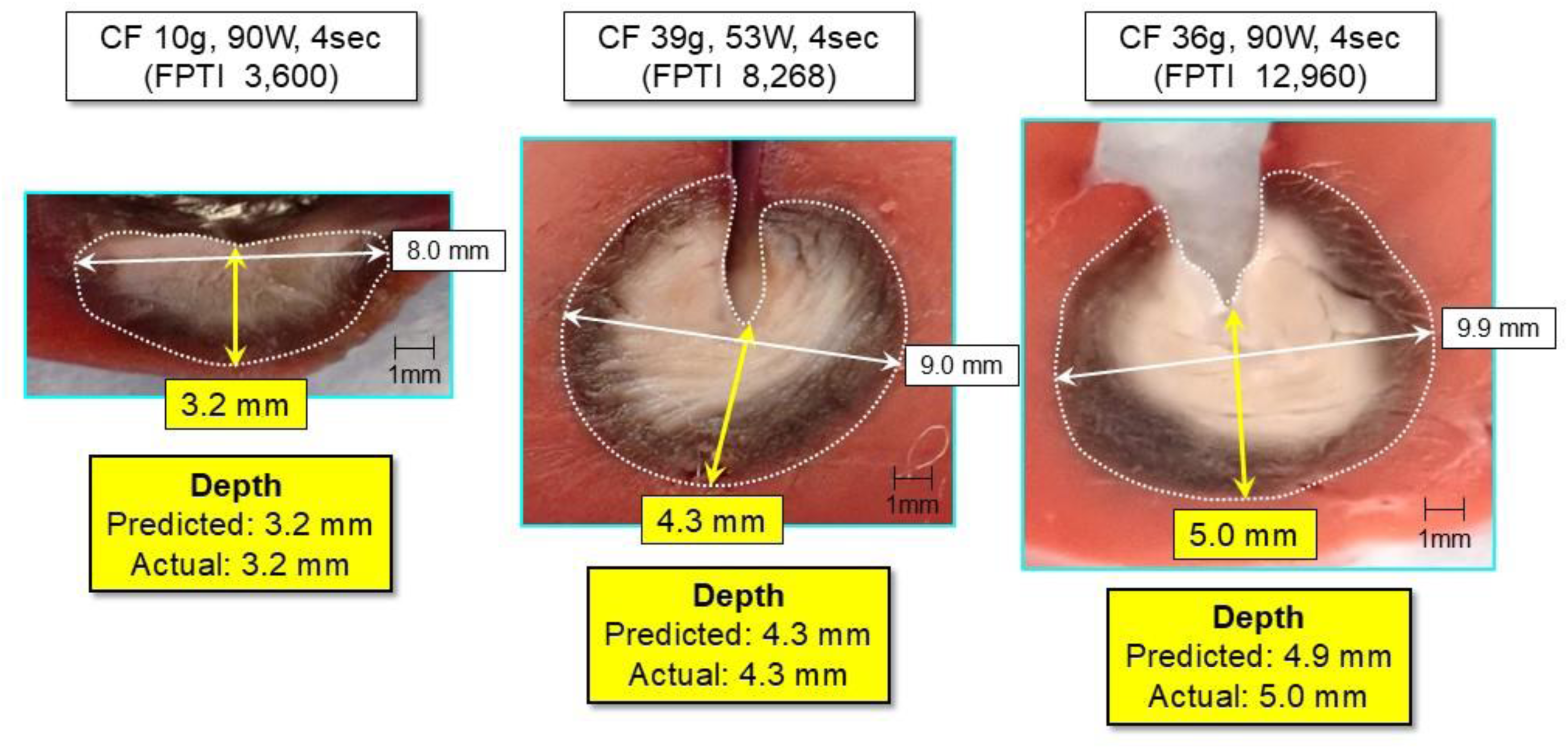
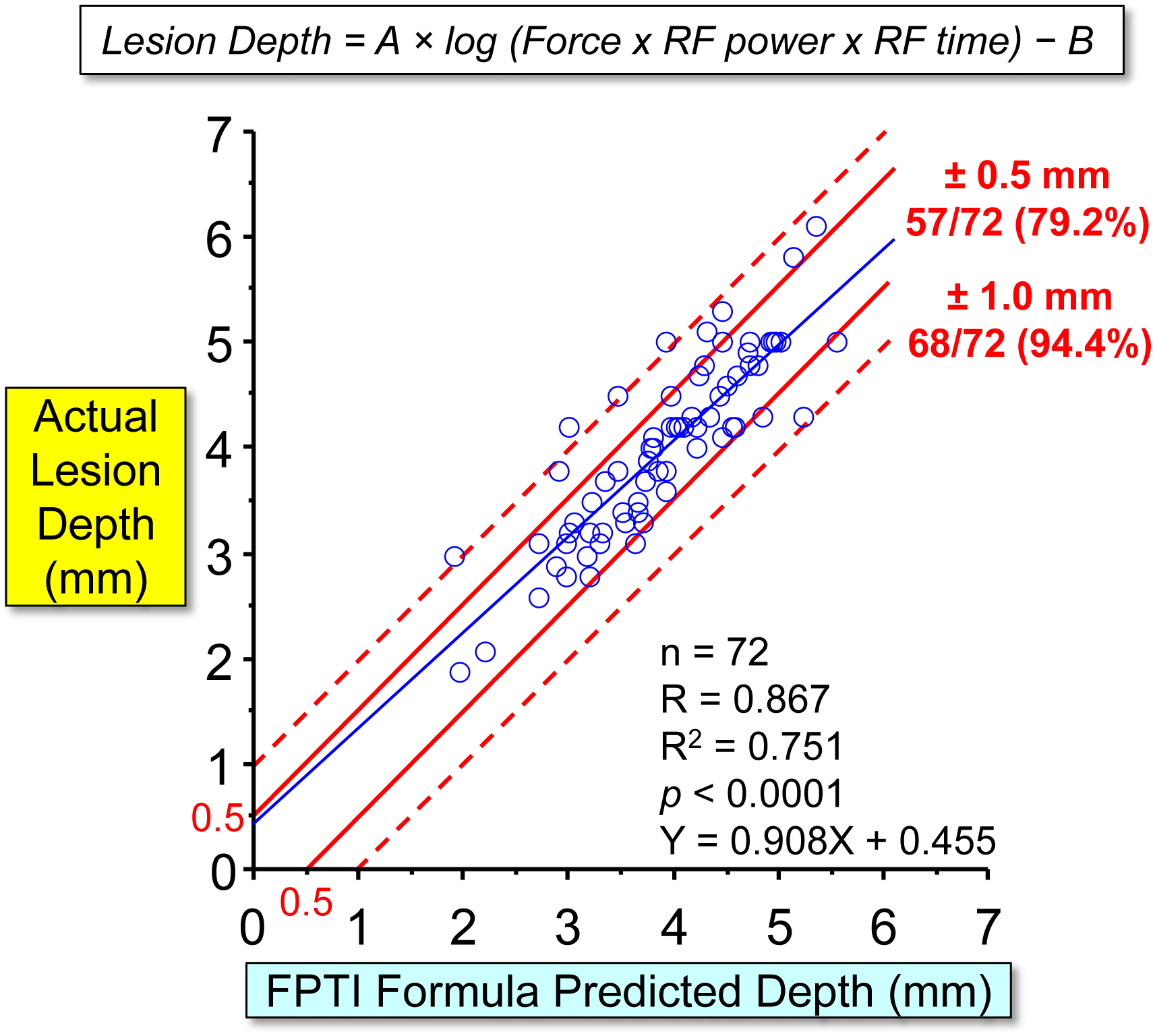
Relationship between the FPTI formula’s predicted lesion depth and actual lesion depth in the prospective validation study. **A.** Cross-section of the ablation lesions to compare the FPTI formula’s predicated lesion depth and actual lesion depth. HP-SD ablation with the predicted lesion depths of 3.2mm (average CF 10*g*, median power 90 Watts for 4sec, FPTI 3,600), 4.3mm (39*g*, 53 Watts for 4sec, FPTI 8,268), and 4.9mm (36*g*, 90 Watts for 4sec, FPTI 12,960) result in actual lesion depths of 3.2mm, 4.3mm, and 5.0mm, respectively, showing the accurate prediction of lesion depth using the FPTI formula. Notice that lower RF power delivery (53 Watts) to maintain the surface electrode temperature of <65°C at the grade 3 trabeculation site (middle panel). **B.** The relationship between the FPTI formula’s predicated lesion depth and actual lesion depth, demonstrating the high accuracy to predict lesion depth (R=0.867, R^2^=0.751, *p*<0.0001, Y=0.908X + 0.455), with ±0.5 mm prediction accuracy in 57/72 (79.2%) lesions and ±1.0 mm prediction accuracy in 68/72 (94.4%) lesions.

#### 3) Data Analysis for Prospective Validation of FPI-Formula to Predict Lesion Depth

The relationship between the FPTI-Formula’s predicted lesion depth and actual lesion depth was compared in each of the 72 lesions (**Fig 8B**).

### Statistical Analysis

Statistical analyses were performed using Stat View software version 5.0 and JMP software version 16.2.0 (SAS Institute, Inc., Cary, NC). Continuous variables are presented as mean ± SD for normally distributed variables. Median with range and interquartile range (IQR) are also shown for non-normally distributed variables. Lesion size (maximum depth, maximum diameter, surface diameter, depth at maximum diameter, and lesion volume) was compared between the three levels of CF using Fisher’s PLSD test or Scheffe test, as appropriate. Comparison of lesion size between with and without RF power modulation was also undertaken with the student *T*-test or Mann-Whitney *U*-test, as appropriate. Categorical variables are described as absolute values and percentages. The comparison between categorical variables was performed with the χ^2^ test or the Fisher exact test, as indicated. The correlation between lesion size (maximum depth and diameter) and each ablation parameter (average CF, median RF power, impedance decrease, and surface electrode temperature) was assessed by the univariable logarithmic regression analysis. The univariable logarithmic regression analysis also assessed the relationship between lesion size and FPTI value. The relationship between FPTI-Formula’s predicted lesion depth and actual lesion depth was assessed by linear regression analysis in phase 1 and 2 studies. The significance of the correlation was determined based on Pearson’s correlation coefficient (R-value) and coefficient of determination (R^2^ value). A probability value (*p*-value) of <0.05 was considered to be statistically significant.

## RESULTS

### Phase 1 Study

A total of 105 RF applications (54 in LV and 51 in RV) were delivered in 5 swine at three levels of CF: 1) low average CF (range 5 to 15*g*, median 10*g*, n=36); 2) moderate average CF (range 16 to 30*g*, median 22*g*, n=35); and 3) high average CF (31-54*g*, median 40*g*, n=34, **Fig 2A**). Neither steam pop nor impedance rise (defined as >10 Ohm increase from the minimal impedance value) occurred during ablation. After ablation, no thrombus was observed on the ablation electrode.

After slicing the TTC-stained ventricular myocardium, all 105 ablation lesions (54 LV and 51 RV lesions) were clearly identified macroscopically (**Fig 3A**). There was no thrombus or crater formation on the endocardium. One hundred-three of the 105 lesions (98%) were non-transmural, while the remaining two lesions were transmural, one observed in the LV apex (average CF 26*g* and median RF power of 90 Watts) and one in the RV apex (34*g* and 86 Watts). Lesion size measurement was excluded in these two transmural lesions because the measurement would be artificially low. Therefore, a total of 103 lesions (53 LV and 50 RV) were included in the following data analysis.

### Relationship Between CF, RF Power, Impedance Decrease, Surface Electrode Temperature, and Lesion Size

In the 103 HP-SD RF applications, RF power was modulated to an average of 77.0±12.9 Watts (range 41-90 Watts) to maintain the surface electrode temperature of <65°C (**Fig 2B** and **Supplemental Table 1**). During HP-SD ablation over the wide range of average CF (5-54*g*, median 22*g*), there is no significant relationship between median RF power and lesion depth (R=0.053, *p*=0.2482) and diameter (R=0.210, *p*=0.3831), respectively (**Fig 4A**). In contrast, there were significant correlations between average CF and lesion depth (R=0.711, *p*<0.0001) and diameter (R=0.430, *p*<0.0001), respectively (**Fig 4B**).

Compared to RF applications with significant power modulation (defined as median power of <80 Watts, average 65.6±8.6 Watts, range of 41-79 Watts, n=50) to maintain the surface electrode temperature of <65°C, RF applications with full power applications (defined as median power of 80-90 Watts, average 87.9±3.0 Watts, n=53) resulted in significantly greater lesion size (maximum depth, maximum diameter, surface diameter, and lesion volume) at moderate and high CF and a trend of greater lesion size (maximum depth, maximum diameter, surface diameter, and lesion volume) at low CF (**Fig 3B** and **Supplemental Table 1**).

Compared to moderate and high CF, low CF ablation was associated with significantly smaller lesion size (maximum depth, maximum diameter, and lesion volume) for both the full and modulated RF power groups (**Fig 3B**). The surface diameter was significantly smaller with low CF compared to moderate CF for the full RF power applications.

The ratio of maximum diameter to maximum depth (**Ratio of Diameter/Depth**) was greatest for low CF, followed by moderate CF, and smallest for high CF for the full and modulated RF power groups (**Fig 3B**).

During HP-SD ablation, the impedance decrease ranged from 9-26 Ohm, with a median of 18 (IQR 4) Ohm. There were weak correlations between the degree of impedance decrease and lesion depth (R=0.312, *p*=0.0013) and diameter (R=0.203, *p*=0.0396), respectively (**Fig 4C, upper panels**). There was a weak correlation between the surface electrode temperature and lesion depth (R=0.272, *p*=0.0357) and no correlation with diameter (R=0.010, *p*=0.1955) (**Fig 4C, lower panels**).

As the trabeculation grade increased at the ablation site, the frequency of RF power modulation to maintain the surface electrode temperature of <65°C significantly increased (17% for grade 1, 62% for grade 2, and 88% for grade 3 trabeculation, **Fig 5A**). Although the average CF was similar among the three trabeculation grades, the median RF power significantly decreased as the trabeculation grade increased: 89 Watts for grade 1, 76.5 Watts for grade 2, and 65 Watts for grade 3, *p*<0.01, respectively (**Fig 5B**). Ablation at sites without apparent trabeculation (grade 1) showed significantly lower initial impedance, compared to sites with grades 2 and 3 trabeculations (a median of 103 Ohms vs. 112.5 Ohms and 112 Ohms, respectively, **Fig 5B**).

### Creation of Force-Power-Time Index (FPTI) Formula to Predict Lesion Size

To estimate the amount of RF energy delivered to the myocardium, a product of average CF (gram) x median power (Watt) x application time (sec) was calculated with each RF application (Force-Power-Time Index: FPTI), ranging from 1,788 to 16,764 (median 6,804) in 103 RF applications. FPTI values correlated well with lesion depth (ranging 1.8-5.9 mm, R=0.792, *p*<0.0001) and diameter (ranging 5.2-10.3 mm, R=0.534, *p*<0.0001, **Fig 6A**). The logarithmic formulas based on these relationships were created:

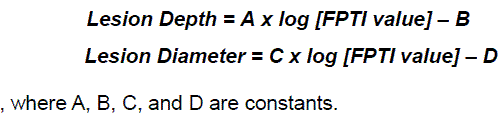

Using these formulas, the difference between the estimated lesion depth and actual lesion depth was within ±0.5 mm in 78/103 (75.7%) lesions and within ±1.0 mm in 97/103 (94.2%) lesions (**Fig 6A**). The difference between the estimated lesion diameter and actual diameter was within ±0.5 mm in 30/103 (29.1%) lesions and within ±1.0 mm in 80/103 (77.7%) lesions (**Fig 6B**).

The FPTI-Formula for the lesion depth was utilized in the phase 2 study to prospectively predict lesion depth during HP-SD ablation (see below).

### Phase 2 Study

In the remaining three swine, HP-SD ablation was performed at 72 separate sites (36 in LV and 36 in RV) with the FPTI-Formula’s predicted lesion depth of 1.9 – 5.6 mm using a wide range of average CF (4-56*g*, median 25.5*g*, **Fig 7**). RF power was modulated to an average of 71.0±18.0 Watts (34-90 Watts) to maintain the surface electrode temperature of <65°C, and the FPTI values ranged from 1,400 to 19,800. There was no steam pop or impedance rise in any of the 72 RF applications. After ablation, no thrombus was present on the ablation electrode or the endocardium at any ablation sites. All 72 ablation lesions were clearly identified macroscopically and non-transmural (**Fig 8A**).

### Prospective Validation of the Accuracy of FPTI-Formula to Predict Lesion Depth

The FPTI-Formula’s predicted lesion depth correlated highly with actual lesion depth (range 1.9 – 6.1 mm) with ±0.5 mm accuracy in 57/72 (79.2%) lesions and ±1.0 mm accuracy in 68/72 (94.4%) lesions (R=0.867, *p*<0.0001) (**Fig 8B**).

## DISCUSSION

The present study demonstrated in a beating heart swine model that, when delivering very high-power RF (≤90 Watts) for very short duration (4 sec), with appropriate power modulation to maintain the surface electrode temperature of <65°C: 1) increasing CF (5-54g) significantly increased lesion depth (1.8-5.9 mm) and diameter (5.2-10.3 mm); 2) lesion depth correlated highly with the wide range of product of average CF (gram) x median RF Power (Watt) x Time (sec) [Force-Power-Time Index, FPTI]; 3) the novel logarithmic FPTI-Formula’s predicted lesion depth correlated with actual lesion depth (1.9 – 6.1 mm) with high accuracy in the prospective validation study, with ±0.5 mm prediction accuracy in 57/72 (79.2%) lesions and ±1.0 mm prediction accuracy in 68/72 (94.4%) lesions (R=0.867, *p*<0.0001); 4) RF power delivery significantly decreased to maintain the surface electrode temperature of <65°C at sites with greater trabeculation (greater electrode-tissue contact area and therefore greater RF power delivery into the tissue), preventing impedance rise, steam pop, and thrombus formation; and 5) there was no or poor correlation between impedance decrease and lesion size or between surface electrode temperature and lesion size.

### Relationship Between RF Parameters and Lesion Size and Creation of a Novel Force-Power-Time Index Formula to Predict Lesion Size During HP-SD Ablation

RF energy delivered into the tissue is converted to thermal energy, resulting in tissue heating and lesion formation. ^11,15,21^ Increased electrode-tissue coupling by increasing CF results in greater RF energy delivered into the tissue, leading to greater tissue heating and lesion formation. ^11–16,21–24^

In the present study, using HP-SD (90 Watts/4 sec) RF applications combined with RF power modulation to maintain the surface electrode temperature of <65°C, increasing CF significantly increased lesion depth and diameter (**Fig 4B**). In contrast, when CF was significantly varied during RF ablation (average CF 5-54*g*), no significant relationship was observed between median RF power and lesion size (**Fig 4A**). RF power recorded from the RF generator represents the total power delivered to the entire ablation circuit (overall power).^25^ However, the proportion of the RF power effectively delivered to the myocardium varies with changes in CF, resulting in significant variation in lesion size even with the same overall RF power delivery.^12–16,21^

To estimate the amount of RF energy delivered to the myocardium, a product of average CF (gram) x median RF power (Watt) x application time (sec) was calculated (Force-Power-Time Index, FPTI). FPTI values correlated well with lesion depth (R=0.792, *p*<0.0001, left panel in **Fig 6A**), suggesting that the FPTI value accurately estimates the RF energy effectively delivered into the myocardium. The logarithmic FPTI formula based on the relationship between lesion depth and FPTI value was created, providing the close relationship between the formula’s calculated lesion depth and actual lesion depth (within ±0.5 mm difference in 78/103 (75.7%) lesions and within ±1.0 mm difference in 97/103 (94.2%) lesions (left panel in **Fig 6B**).

Compared to the correlation between the FPTI value and lesion depth, the correlation between the FPTI value and lesion diameter is relatively lower (R=0.792 vs. R=0.536, **Fig 6B**). Changes in ablation electrode orientation (perpendicular vs. parallel) may result in greater variation of lesion diameter. It has been suggested that greater RF current distribution and lesion formation occur when the ablation electrodes are positioned parallel to the muscle fiber orientation compared with perpendicular to the muscle fiber.^26,27^ The dependency on muscle fiber orientation during RF ablation may also result in greater variation in lesion diameter.

During HP-SD ablation, the impedance decreased from 9 to 26 Ohms, resulting from tissue heating (i.e., a greater decrease in impedance with higher tissue temperature).^15^ However, there was only a poor or no correlation between the impedance decrease and lesion depth and diameter, respectively, limiting the clinical implications of the impedance decrease in predicting lesion size (**Fig 4C**).

### Prospective Validation of the FPTI-Formula to Predict Leion Depth

To our knowledge, this is the first study to prospectively validate the accuracy of the FPTI-Formula for HP-SD (90Watts/4 sec) ablation to predict lesion depth in a beating heart model. The FPTI-formula’s predicted lesion depth with FPTI values of 1,400 – 19,800 (median 6,110), comprised of CF of 4-56*g* (median 25.5*g*) and median RF power of 34-90 Watts (median 76 Watts), correlated very well with actual lesion depth (1.9 – 6.1 mm, median 4.1 mm) with ±0.5 mm accuracy in 57/72 (79.2%) lesions and ±1.0 mm accuracy in 68/72 (94.4%) lesions (R=0.867, *p*<0.0001, **Fig 8B**), without the occurrence of an impedance rise, steam pop or thrombus formation.

In clinical PV isolation studies in AF patients using the conventional RF Ablation Index with RF conventional RF power (25-45 Watts) and duration (15-60 sec), target Ablation Index values of 350-400 (predicted lesion depth of 3.5-4.0 mm) for the posterior/inferior left atrial wall and 500-600 (predicted lesion depth of 5.0-6.0 mm) for the anterior/superior wall have been utilized, resulting in a high incidence of complete ipsilateral PV isolation with the first encirclement and high durability of PV isolation.^17–20^ The present study demonstrates that the novel FPTI-Formula for HP-SD ablation can accurately predict a similar range of lesion depth (2-6mm), suggesting its potential to facilitate PV isolation by creating transmural, continuous atrial lesions with shorter RF times.

### Relationship Between Surface Electrode Temperature and the Myocardial Trabeculation Grade

Greater reduction in RF power was required (power modulation) to maintain the surface electrode temperature of <65°C at sites with greater myocardial trabeculation (**Fig 5**). When the ablation catheter is positioned in a deep trabecular groove, even with low CF, the electrode-tissue coupling area may increase, allowing a greater proportion of RF energy to be delivered to the myocardium, leading to greater myocardial heating. The higher initial impedance at sites with greater trabeculation also indicates greater electrode-tissue coupling (**Fig 5B**). In this condition, without power modulation, high-power RF delivery may increase the risk of overheating within the tissue and at the electrode-tissue interface, potentially causing steam pops and thrombus formation. In addition, electrode temperature during saline irrigation becomes less reflective of tissue temperature, increasing the risk of steam pops due to tissue overheating. ^28,29^ The Qdot catheter used in the present study measures the surface electrode temperature with six thermocouples located only 75μm from the electrode surface during a low irrigation rate of 8 ml/min, potentially reducing the discrepancy between electrode temperature and tissue temperature. The surface electrode temperature control by RF power modulation appears to reduce the risk of overheating within the tissue and at the interface, as no impedance rise, steam pops, or thrombus formation was observed in the present study.

### Clinical Implications

The creation of continuous transmural atrial lesions without collateral injury is desired for PV isolation procedures. However, recent clinical studies on PV isolation using 90 Watts/4 sec RF applications have demonstrated significant variability in its efficacy, with first-pass PV isolation rates ranging from 18% to 83% and acute reconnection rates ranging from 4% to 32%.^4–10^ Esophageal injury has also been reported. ^4,30^ In contrast, Ablation Index-guided PV isolation procedures using conventional RF power and longer times have shown high first-pass PV isolation rates, low acute PV reconnection rates, and high lesion durability.^17–20^

In previous studies on 90 Watts/4 sec RF ablation, it has been suggested that the majority of the lesion is produced by resistive heating, which may result in wider but shallower lesions, potentially increasing the efficacy of PV isolation with shorter procedure times and reducing the risk of collateral injury.^1–3^ However, a recent study on 90 Watts/4 sec RF applications has demonstrated that a significant portion of effective tissue heating and lesion formation occurred after RF termination due to slower conductive tissue heating (i.e., thermal latency).^21^ Lower energy of 360 Joules with 90 Watts/4 sec RF delivery is associated with smaller lesion formation (depth and diameter), and lesion size significantly increases with increasing CF. ^21, 31,32^

Using the novel FPTI-Formula developed in the present study, it is feasible to accurately predict lesion depth in real-time during HP-SD ablation. Lesion diameter may also be estimated based on the diameter-to-depth ratio to select the optimal inter-lesion distance. The lesion diameter would be approximately 1.61-1.72 times the predicted lesion depth with high CF, 1.79-1.96 times with moderated CF, and 2.26-2.30 times with low CF (**Fig 3B**). Therefore, HP-SD ablation guided by the novel FPTI-Formula may facilitate the creation of continuous transmural lesions with shorter RF times while preventing collateral injury.

### Study Limitations

One potential limitation of the present study is that HP-SD ablation was performed in the LV and RV and not in the atrium, to allow an accurate measurement of lesion depth by having non-transmural lesions.

Although there was no steam pop or thrombus formation during HP-SD RF applications (90 Watts/4 sec) in the present study, longer application times and higher CF may result in steam pop or thrombus formation.

In the present study, during HP-SD ablation, the average CF with a target depth of 3-4 mm is 15*g*, while higher CF (average 34*g*) is required with a target depth of 5-6 mm (**Supplemental Fig 1**). When ablation is performed at a thicker atrial wall (such as the narrow left anterior ridge),^33,34^ it may be challenging to achieve stable high CF. In such cases, switching to the conventional Ablation Index-guided ablation may be required using lower CF, moderate power, and a longer RF application time.^9,17^

Finally, clinical studies are required to determine the efficacy and safety of HP-SD ablation guided by the novel FPTI-Formula, including a first-pass PV isolation rate, lesion durability, and the potential reduction of the risk of collateral injury.

## CONCLUSIONS

During HP-SD (90 Watts/4 sec) RF ablation in a closed chest beating heart swine model, lesion depth correlated highly with the product of average CF (gram) x median RF Power (Watt) x Time (sec) [Force-Power-Time Index, FPTI]. In this prospective validation study, a novel logarithmic FPTI-Formula’s predicted lesion depth correlated well with actual lesion depth (1.9 – 6.1 mm), with ±0.5 mm prediction accuracy in 57/72 (79.2%) lesions and ±1.0 mm prediction accuracy in 68/72 (94.4%) lesions (R=0.867, *p*<0.0001). Significant RF power modulation to maintain the surface electrode temperature of <65°C at sites with greater trabeculation reduced the risk of impedance rise, steam pop, and thrombus formation.

### Sources of Funding

This study was supported, in part, by a grant from Johnson & Johnson MedTech,

Inc.

### Disclosures

Dr. Nakagawa is a consultant for Johnson & Johnson MedTech, Inc., Abbott, Inc., CardioFocus, Inc, Stereotaxis, Inc, Japan Lifeline, Ltd, Fukuda Denshi, Ltd and Philips, Japan, Inc. Dr. Jackman is a consultant for Johnson & Johnson MedTech, Inc. Drs. Govari, Bubar, Sharma, and Beeckler are employees of Johnson & Johnson MedTech, Inc. The other authors report no conflicts.

### Supplemental Materials

Supplemental Table 1 and Figure 1.

## Nonstandard Abbreviations and Acronyms

PV: pulmonary vein
AF: atrial fibrillation
RF: radiofrequency
CF: contact force
ICE: intracardiac echocardiography
HP-SD: high-power and short-duration
FPTI: Force-Power-Time Index
LA: left atrium
LV: left ventricle
RV: right ventricle

